# COVID-19 vaccine distrust in Colombian university students: Frequency and associated variables

**DOI:** 10.1101/2021.03.07.21253080

**Authors:** Adalberto Campo-Arias, John Carlos Pedrozo-Pupo

## Abstract

The study aimed to know the frequency and variables associated with COVID-19 vaccine distrust in students of a Colombian university. A cross-sectional study was carried out which participated emerging adult students of a Colombian university. A total of 1,136 students between 18 and 29 years (M= 22.0, SD = 3.0); most of them were female (66.0%), non-health students (82.8%), low-income (79.0%), and residents of urban areas (84.9%). It was frequent low institutional trust (74.8%), low cognitive, social capital (27.9%), low fear of COVID-19 (49.5%), low perceived stress related to COVID-19 (83.5%), and high COVID-19 vaccine distrust (78.9%). Non-health carrier (Adjusted OR = 3.63, 95%CI 2.58-5.10), rural residence (AOR = 1.85, 95%CI 1.13-3.04), low income (AOR = 1.84, 95%CI 1.31-2.57), and perceived stress related to COVID-19 (AOR = 1.74, 95%CI 1.20-2.54) were related to high COVID-19 vaccine distrust. In conclusion, COVID-19 vaccine distrust is high among emerging adult Colombian university students. The COVID-19 vaccine distrust is related to non-health science carriers, rural residents, low-income, and low-perceived stress related to COVID-19. The COVID-19 related health literacy should be improved in students of this university considering socio-cultural background.

## Introduction

In Colombia, the first case of coronavirus disease (COVID-19) was identified on March 6th, 2020; since one year of the pandemic, more than 2.2 million cases and 60 thousand deaths from COVID-19 have been confirmed, 1,177 per million inhabitants [1]. However, mistrust towards COVID-19 vaccines has been evidenced in national news and social networks [2], as activism against some vaccines or national vaccination plans has increased [3, 4].

According to the World Health Organization, the spectrum of vaccination intentions to receive a COVID-19 vaccine follows a non-linear pattern that includes activist, rejecting, hesitance, accepting, demanding, and advocating. The opposing faces, activism implies active participation in events such as protests; rejection is based on the safety of COVID-19 vaccines, experiences, perceptions, and values; and hesitation is the presence of doubts in accepting the COVID-19 vaccine due to the novelty of the disease, ignorance of the vaccination program, use of new vaccination platforms and uncertainty surrounding the safety of the vaccine [5]. Given the overlap in these definitions, they are often referred to interchangeably as aversion, distrust, hesitance, mistrust, refusal, rejection, or resistance [3, 4].

The frequency of distrust towards the COVID-19 vaccine varies widely depending on the context, way of measurement, and pandemic stage [6-8]. Sallam, among adults representing the general public, found prevalences between 3% in Ecuador and 76.4% of vaccine distrust in Kuwait; 5.7% in Malaysia, 6.7% in Indonesia, 8.7% in China, 41.1% in France, 43.1% in the United States, 43.7% in Poland, 45.1% in Russia, 46,3% in Italy, and 71.6% in Jordan [9].

Similar findings have been observed in college or university students. For instance, Barello et al. [10], among 735 Italian students, 86.1% reported intention to get vaccinated, and 13.9% reported not being sure [low intention to vaccinate]. In Malta, Grech et al. [11] found that in 852 students, 30.5% reported unlikely, and 25.3% undecided to receive the COVID-19 vaccine.

The variables associated with distrust of COVID-19 vaccines follow the same pattern as the markedly different frequencies in the available studies; it highlights the importance of cultural and social determinants of health literacy [12]. Barello et al. [10] found no demographic differences between university students who accept vaccination and those who have low intention. Grech et al. [11] reported that COVD-19 vaccine distrust was higher in dentistry than medical students and university administrative personnel than academic and students. However, among the general population, Fisher et al. [13] observed that distrust of COVID vaccines was associated with younger age, Afro-American, lower educational achievement, and not receiving the influenza vaccine in the last year. Paul et al. [14] reported COVID-19 vaccine uncertainty and refusal were associated with female gender, low-income, having not received a flu vaccine prior year, poor adherence to COVID-19 government guidelines, and living with children among a large sample from the general population.

Few studies have quantified attitude using construct measurement scales, none in a low- or middle-income country [9]. Careful consideration should be given to the need for specific public health actions according to university students’ characteristics since health knowledge can have a lasting positive impact on the course of life [15]. Besides, university students adequately instructed in scientific information on the COVID-19 pandemic can be disseminators, promoters, or influencers of vaccination among peers and relatives with less access to truthful information [16].

The study’s objective was to evaluate the frequency and variables associated with COVID-19 vaccine distrust in students of a Colombian university.

## Methods

### Participants

An analytical cross-sectional study was implemented. Non-probability sampling was taken. At least 384 participants were expected; this sample size is adequate for prevalence from 3% (more or less 1) to 50% (more or less 5) for any variable, a confidence level of 95% [17]. This sample is indicated to explore associations between variables with sufficiently narrow confidence intervals. 21.114 emails were sent to active pre-graduate students. It is expected to have students of an emerging age, that is, between 18 and 29 years old [18]. Since the mailing list provided omitted demographic characteristics, the email was sent regardless of age. However, the questionnaire’s presentation only invited participants from the age of 18 because minors’ participation requires parents’ or guardians’ consent in Colombia.

### Measurements

#### COVID-19 Vaccine Attitude Scale

The Spanish version of the COVID-19 Vaccine Attitude Scale (COVID-19 VAS) is a translation process from the original version in Danish to English and English to Spanish. The back-translation process was satisfactory because the sentences’ simplicity easily made it possible to reach equivalence. The COVID-19 VAS is an eight-item tool that offers five response options: “totally disagree”, “disagree”, “unsure”, “agree”, and “totally in agreement”. These responses are scored directly from 0 to 4, except for item 7, which is scored in reverse. The possible total scores are between 0 and 32. The scale presents high internal consistency, Cronbach’s alpha of 0.95 [19]. Scores of 24 or less were classified as high COVID vaccine distrust. It was taken in the third quartile as the cut-off point. In the present sample, internal consistency was 0.95

#### Institutional trust

Institutional trust was measured with an instrument adapted for the COVID-19 outbreak, the Institutional Trust Scale (ITS) [20]. The ITS is an instrument made up of four items that ask for trust in the national government and the municipality (mayors). Each item offers four response options: “strongly disagree,” “disagree,” “agree,” and “strongly agree,” which are scored from 0 to 3. The ITS was used in a previous study for exploitative purposes without reporting reliability or validity indicators; however, it showed acceptable face validity [19]. In the present study, scores between 0 and 5 were classified with low institutional trust. The internal consistency of the ITS was 0.62 in the current sample.

### Cognitive capital social

The cognitive social capital scale (CSCS) measures global social capital and explores social cohesion and trust in the neighborhood and nearby community. The CSCS presents five questions with Likert response options: “very disagree,” “disagree,” “agree,” and “very agree,” that rated from 0 to 3. The original study did not report internal consistency (21). The CSCS showed Cronbach’s alpha value of 0.79 in a Colombia sample [22]. In the present investigation, scores less than or equal to five were categorized as low CSC. Internal consistency of 0.81.

#### COVID-5 Fear Scale (Fear of COVID-5)

The Fear of COVID-5 is a five-item tool that quantifies attitude toward COVID-19. The scale offers four response options: “never,” “seldom,” “often,” and “always,” which are rated from zero to three. Total scores are between 0 and 15; scores lower than four were classified as low fear of COVID-19. The Fear of COVID-5 showed an internal consistency of 0.75 in Colombian adults who participated in an online investigation [23]. The Fear of COVID-5 presented a Cronbach’s alpha of 0.82 in the current sample.

#### COVID-19 Pandemic-Related Perceived Stress Scale (PSS-10-C)

The PSS-10-C is a ten-item instrument that quantifies perceived stress during the last month related to the COVID-19 epidemic. The tool offers five response options: “never,” “seldom,” “occasionally,” “often,” and “always,” which are rated from zero to four. The total scores can be observed between 0 and 40; scores below 25 were categorized as low perceived stress. The PSS-10-C showed an internal consistency of 0.85 in a previous online study of Colombian adults [24]. The PSS-10-C reached internal consistency of 0.85.

### Procedure

An invitation was sent from one of the researchers’ institutional mail to the mail consigned in the university’s admission system. The email explained the study’s objectives and reported the participants’ anonymity and the confidential handling of information. Besides, informed consent was required to implement a questionnaire designed by Google Form^(c)^. All responses were mandatory to avoid missing values. Two emails were sent with an interval of one week. In the second, those who had not done so were recalled and invited to participate. A second email a week can improve the response rate [25]. The questionnaire link was active for prospective participants between January 18th and February 5th, 2021.

### Data analysis

For qualitative variables, frequencies (n) and percentages (%) were established, and for quantitative variables, mean (M), standard deviation (SD), median, and interquartile range (IQR). Variables were dichotomized for bivariate analysis to avoid the excess of categories, with the danger of observing only associations that are not statistically significant. These no associations could be false; that is, it avoids making a type II statistical error. The COVID-19 vaccine distrust was the dependent variable, demographic features, institutional trust, cognitive social capital, fear of COVID-19, and stress-related to COVID-19, as independent factors. Crude and adjusted odds ratios (OR) were established with 95% confidence intervals (CI).

The associations were adjusted according to Greenland’s recommendations for logistic regression [26]. The first recommendation is that all variables that show significant association or probability values less than 0.20 be considered for the adjustment. Furthermore, the second is that only the variables that show significant probability values are left in the final model, and those variables that induce a modification of 10% or more in the most robust association, in the process it is placed in the first order. Those non-significant variables that produce a variation of 10% or more indicate a significant confounding effect that warrants control. The Hosmer-Lemeshow goodness-of-fit test was computed for each adjustment [27]. Statistical analysis was performed with IBM-SPSS version 23.0 [28].

### Ethical issues

The project respected the ethical principles promulgated in Resolution 008430 of 1993 of the Colombian Ministry of Health and Declaration of Helsinki of the World Medical Association. An institutional research ethics board approved the project (Act 002 of the ordinary meeting of March 26th, 2020). All participants must sign an online informed consent to participate.

## Results

A total of 1,409 (6,7%) out of 21,144 pre-graduate students responded to the email. Two hundred seventy-three questionnaires were excluded because responders were not emerging adults; 67 students under 18 and 206 over 29 years. Then, 1,136 questionnaires of emerging adult students (M=22.0, DE=3.0) were analyzed.

The highest proportion of responders were women, single, non-health students, low-income, and residents of urban areas. The characteristics of the participants and other variables are presented in Table 1.

**Table 1.**
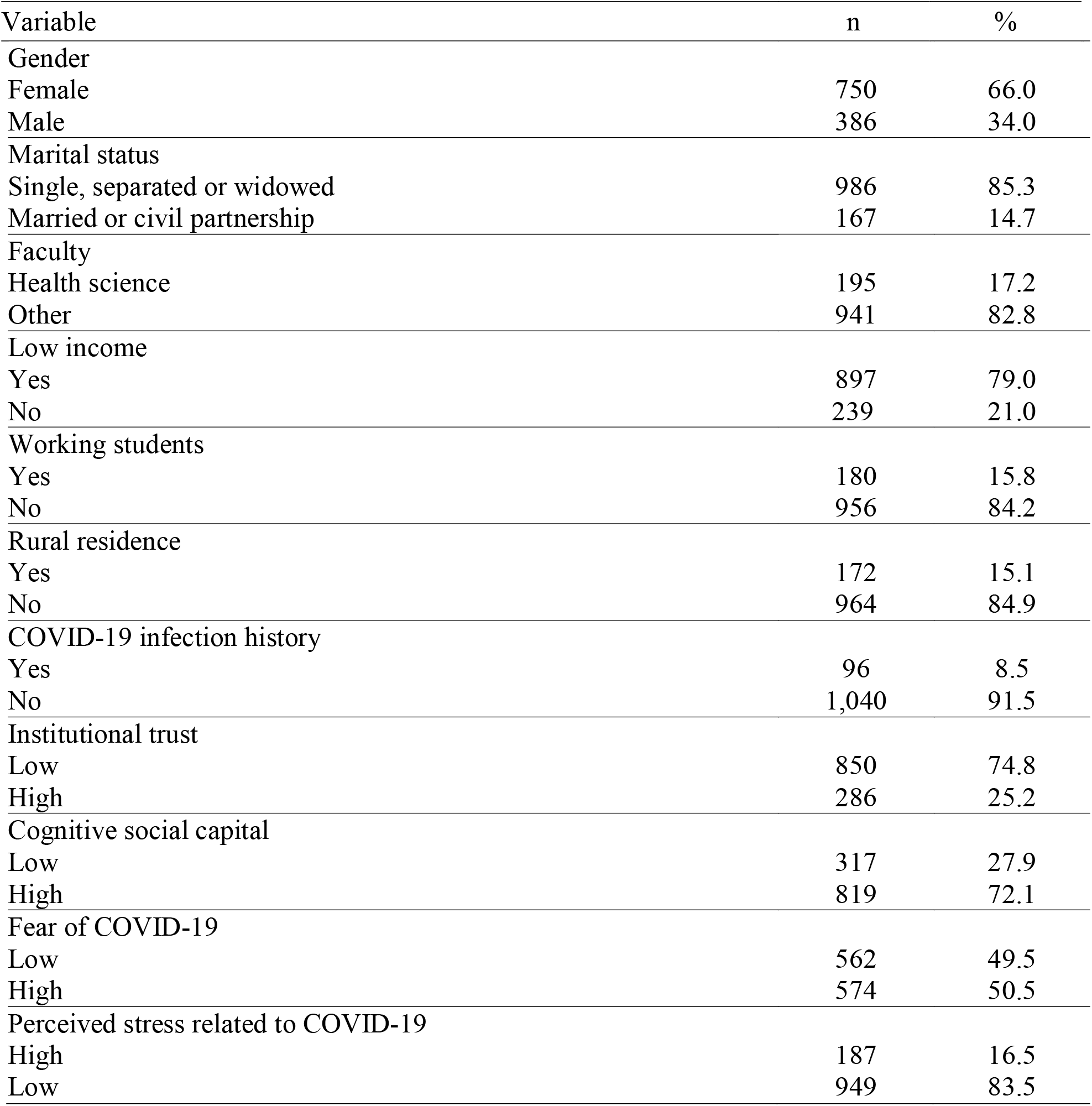
Demographical characteristics of sample.

The COVID-19 VAS scores were found between 0 and 32 (M=18.7); 78.9% (n = 896) students reported high; and 21.1% (n = 240), low COVID-19 vaccine distrust (or high trust in COVID-19 vaccination). More scores on the measurement scales are presented in Table 2. Crude associations were observed between the carrier, residence place, income, marital status, perceived stress related to COVID-19, and high COVID-19 vaccine distrust. Associations are presented in Table 3. Nevertheless, marital status was not associated with high COVID-19 vaccine distrust after adjusting for the other variables, and it did not affect the final model that showed adequate goodness-of-fit. See details in Table 4.

**Table 2.** Scores on the scales.

| Scale | Range | M (SD) | Me (IQR) |
| --- | --- | --- | --- |
| Institutional Trust Scale | 0-12 | 4.3 (2.1) | 4 (3-6) |
| Cognitive Social Capital Scale | 0-15 | 7.2 (3.0) | 7 (5-9) |
| Fear of COVID-19 | 0-15 | 5.0 (4.0) | 3 (2-5) |
| PSS-10-C | 0-40 | 18.2 (4.4) | 19 (14-22) |
| COVID-19 VAS | 0-32 | 18.7 (7.2) | 19 (14-24) |

**Table 3.** Crude association for COVID-19 vaccine distrust.

| Variable | OR | 95%CI |
| --- | --- | --- |
| Female gender | 1.16 | 0.86-1.56 |
| Married or civil partnership | 1.62 | 1.03-2.54 |
| Non-health science student | 3.50 | 2.51-4.87 |
| Low income | 1.97 | 1.43-2.73 |
| Working student | 1.23 | 0.82-1.85 |
| Rural residence | 2.11 | 1.31-3.42 |
| Non history of COVID-19 | 0.85 | 0.50-1.45 |
| Institutional distrust | 0.99 | 0.71-1.37 |
| Low cognitive social capital | 1.00 | 0.73-1.37 |
| Low fear of COVID-19<sup>†</sup> | 1.01 | 0.76-1.34 |
| Low perceived stress related to COVID-19 | 1.51 | 1.05-2.16 |

**Table 4.** Adjusted association for COVID-19 vaccine distrust.

| Variable | OR | 95%CI |
| --- | --- | --- |
| Non health science student | 3.63 | 2.58-5.10 |
| Rural residence | 1.85 | 1.13-3.04 |
| Low income | 1.84 | 1.31-2.57 |
| Low perceived stress related to COVID-19 | 1.74 | 1.20-2.54 |

Hosmer-Lemeshow' test, $X^2=1.98$ , $df=5$ , $p=0.85$ .

## Discussion

The frequency of COVID-19 vaccine distrust is 78.9%, and it is related to non-health science carrier, rural residence, low income, low perceived stress related to COVID-19 in a sample of students at a Colombian university. The frequency observed is significantly lower than documented in university students in Italy, in whom 13.9% reported a low intention to vaccinate [10], and in Malta, where 55.8% of participants reported that it was unlikely or undecided about being vaccinated against COVID-19 [11]. These marked differences have also been documented in the general population [6-9].

This population is expected to find a high frequency of trust in COVID vaccines for two reasons: the first, emerging adults were included, and the second, being in higher education carriers. The findings are a strong wake-up call for educational and public health authorities to better understand student health education, particularly related to COVID-19, given the high safety and positive effects of the vaccination [29]. Health literacy is higher in the younger cohorts and people with more formal education years [13, 14, 30]. It is postulated that the ability of discernment, above the average, in university students should be a protective factor against the infodemic associated with COVID-19 [30, 31].

Furthermore, in the present study, it was observed that low income and rural residence were related to high distrust towards COVID-19 vaccination. These findings are consistent with other studies that show that these characteristics predict distrust of vaccines [13, 14]. However, another investigation found independence between income and scores for attitude toward COVID-19 vaccines [32]. However, it should be considered that the attitude towards vaccines, in general, is related to cultural and social factors that can be little affected by professional training [12]. For instance, the current report failed to show an association between institutional trust, social capital, and vaccine distrust. This finding is striking because Colombia is an upper-middle-income country with a low Gini inequality index of 0.54 in 2018 [33] and a high corruption index of 39 in 2020 [34]. In Italy, Prati [35] found institutional trust related to COVID-19 vaccine trust. Public vaccination trust in programs depends on the work institutions do for the community regarding individual, family, cultural, social, and political issues [36].

This preliminary finding suggests that student’s health literacy influences the vaccination attitude. In Malta, it was found that medical students reported higher COVID-19 vaccine trust than dentistry students and administrative workers [11]. Similarly, psychological states such as perceived stress can anticipate the attitude towards vaccines. Previously, it was reported that worries were correlated to COVID-19 vaccine trust [35]. Nevertheless, fear of COVID-19 may have less importance; Paul et al. [14] reported an independent correlation between Coronavirus Anxiety Scale and mistrust of vaccine benefits scores. These associations require further study to get strong evidence and variables such as gender that have shown inconsistent associations [10, 13, 14].

Nowadays, strengthening health literacy is very important to prepare individuals and societies for future emergencies that require urgent actions and rapid inter-and cross-sectoral containment. Educational institutions, social media, work environments, healthcare systems, political sectors, and economic groups play a crucial role to reduce the negative impact of emerging diseases and build individual, family, community, and society health literacy; possibly, for healthy collectives is required more and better health education [12]. The COVID-19 pandemic is more of a syndemic as a result of the complex interaction of negative individual, social and cultural events, processes, and situations that converge in poverty, financial or multidimensional; consequently, people living in poverty are at greater risk of fall ill or die from the convergence of acute and chronic adversities [37, 38].

This study provides relevant information on COVID vaccines’ attitude, measured with a structured instrument, and the importance of psychological distress in emerging-age university students from a middle-income country. Nevertheless, the study has several limitations that invite one to take the findings with caution. The sample does not represent Colombian university students [17], the response rate was relatively low [25], and reliability of the institutional trust measurement showed a value lower than 0.70 [39]; although, depending on the development of the instrument, values greater than 0.60 may be allowed [40].

In conclusion, the frequency of COVID-19 vaccine distrust is high among emerging adult pre-graduate students at a Colombian university. The COVID-19 vaccine distrust is associated with non-health science carriers, rural residents, low-income and low-perceived stress related to COVID-19. COVID-19 related health literacy should be improved in this group of students. All training actions must consider the social and cultural context of the students.

## Data Availability

The data that support the findings of this study are available from the corresponding author upon reasonable request.

## References

1. Ministerio de Salud y la Protección Social de Colombia. [Current situation: new Coronavirus (COVID-19)]. Accessed March 6th, 2021. https://www.minsalud.gov.co/salud/publica/PET/Paginas/Covid-19_copia.aspx.

2. El Tiempo. [Why is there mistrust in vaccines against covid-19?] Accessed January 8th, 2021. https://www.eltiempo.com/mundo/mas-regiones/vacuna-coronavirus-por-que-hay-desconfianza-en-la-vacuna-contra-el-covid-19-559458

3. Dubé E, Vivion M, MacDonald NE. Vaccine hesitancy, vaccine refusal, and the antivaccine movement: influence, impact, and implications. Expert Rev Vaccines. 2015; 14(1): 99–117.

4. Zaidi MB, Flores-Romo L. The growing threat of vaccine resistance: a global crisis. Curr Treat Opt Infect Dis. 2020; 12: 122–134.

5. World Health Organization. Covid-19 vaccines: safety surveillance manual. Geneva: World Health Organization; 2020.

6. Lin C, Tu P, Beitsch LM. Confidence and receptivity for COVID-19 Vaccines: A rapid systematic review. Vaccines. 2021; 9 (1):16.

7. Feleszko W, Lewulis P, Czarnecki A, Waszkiewicz P. Flattening the curve of COVID-19 vaccine rejection—An international overview. Vaccines. 2021; 9 (1): 44.

8. Robinson E, Jones A, Daly M. International estimates of intended uptake and refusal of COVID-19 vaccines: A rapid systematic review and meta-analysis of large nationally representative samples. Vaccine 2021. https://doi.org/10.1016/j.vaccine.2021.02.005

9. Sallam M. COVID-19 vaccine hesitancy worldwide: A concise systematic review of vaccine acceptance rates. Vaccines. 2021; 9 (2): 160.

10. Barello S, Nania T, Dellafiore F, Graffigna G, Caruso R. ’ Vaccine hesitancy among university students in Italy during the COVID-19 pandemic. Eur J Epidemiol. 2020; 35 (8): 781–783.

11. Grech V, Gauci C. Vaccine hesitancy in the University of Malta Faculties of Health Sciences, Dentistry and Medicine vis-a-vis influenza and novel COVID-19 vaccination. Early Hum Dev. 2020; 105258.

12. Abdel-Latif MM. The enigma of health literacy and COVID-19 pandemic. Public Health. 2020; 185: 95–96.

13. Fisher KA, Bloomstone SJ, Walder J, Crawford S, Fouayzi H, Mazor KM. Attitudes toward a potential SARS-CoV-2 vaccine: A survey of US adults. Ann Intern Med. 2020; 173 (12): 964–973.

14. Paul E, Steptoe A, Fancourt D. Attitudes towards vaccines and intention to vaccinate against COVID-19: Implications for public health communications. Lancet Regional Health-Europe, 2020; 1: 100012. https://doi.org/10.1016/j.lanepe.2020.100012

15. Dunne C, Somerset M. Health promotion in university: what do students want? Health Educ. 2004; 104 (6): 360–370.

16. Mendoza-Núñez VM, Mecalco-Herrera C, Ortega-Ávila C, Mecalco-Herrera L, Soto-Espinosa JL, Rodríguez-León MA. A randomized control trial: training program of university students as health promoters. BMC Public Health. 2013; 13 (1): 162.

17. Hernández J. [Size simple for a clinical trial]. Rev Colomb Gastroenterol. 2006; 21 (2): 118–121.

18. Arnett JJ. Emerging adulthood: A theory of development from the late teens through the twenties. Am Psychol. 2000; 55 (5): 469–489.

19. De Roos NI. Who should tell you to vaccinate? the role of the provider of information in the attitude towards the Corona vaccine: Need for conformity as a moderator? (master’s thesis). Utrech: Utrech University; 2020.

20. Prati G, Pietrantoni L, Zani B. Compliance with recommendations for pandemic influenza H1N1 2009: the role of trust and personal beliefs. Health Educ Res. 2011; 26 (5): 761–769.

21. Martin KS, Rogers BL, Cook JT, Joseph HM. Social capital is associated with decreased risk of hunger. Soc Sci Med. 2004; 58 (12): 2645–2654.

22. Caballero-Domínguez CC, De Luque-Salcedo JG, Campo-Arias A. Social capital and psychological distress during Colombian coronavirus disease lockdown. J Community Psychol. 2021; 49 (2): 691–702.

23. Cassiani-Miranda CA, Tirado-Otálvaro AF, Campo-Arias A. Adaptation and psychometric evaluation of the Fear of COVID-19 Scale in the general Colombian population. Death Stud. 2021 (ahead of print). https://doi.org/10.1080/07481187.2021.1874572

24. Campo-Arias A, Pedrozo-Cortés MJ, Pedrozo-Pupo JC. Pandemic-Related Perceived Stress Scale of COVID-19: An exploration of online psychometric performance. Rev Colomb Psiquiatr 2020;49 (4): 229–230.

25. Nulty DD. The adequacy of response rates to online and paper surveys: what can be done? Assess Eval High Educ. 2008; 33 (3): 301–314.

26. Greenland S. Modeling and variable selection in epidemiologic analysis. Am J Public Health. 1989; 79 (3): 340–349. https://doi.org/10.2105/ajph.79.3.340

27. Hosmer DW, Taber S, Lemeshow S. The importance of assessing the fit of logistic regression models: a case study. Am J Public Health. 1991; 81 (12): 1630–1635. https://doi.org/10.2105/ajph.81.12.1630

28. IBM-SPSS Statistics for Windows, version 23.0, 2015. Armonk: SPSS. Inc.

29. Petousis-Harris H. Assessing the safety of COVID-19 vaccines: A primer. Drug Safety. 2020; 43 (12): 1205–1210.

30. McCaffery KJ, Dodd RH, Cvejic E, Ayrek J, Batcup C, Isautier JM, et al. Health literacy and disparities in COVID-19–related knowledge, attitudes, beliefs, and behaviours in Australia. Public Health Res Pract. 2020; 30 (4): 30342012.

31. Carrion-Alvarez D, Tijerina-Salina PX. Fake news in COVID-19: A perspective. Health Prom Perspect. 2020; 10 (4): 290–291.

32. Pogue K, Jensen JL, Stancil CK, Ferguson DG, Hughes SJ, Mello EJ, et al. Influences on attitudes regarding potential COVID-19 vaccination in the United States. Vaccines. 2020; 8 (4): 582.

33. Trading Economy. Colombia Corruption Index. 1995-2020 Data. Accessed March 5th, 2021. https://tradingeconomics.com/colombia/corruption-index

34. World Bank. Gini index (World Bank estimate). Accessed March 5th, 2021. https://data.worldbank.org/indicator/SI.POV.GINI?view=chart

35. Prati G. Intention to receive a vaccine against SARS-CoV-2 in Italy and its association with trust, worry, and beliefs about the origin of the virus. Health Educ Res. 2020; 35 (6): 505–511.

36. Harrison EA, Wu JW. Vaccine confidence in the time of COVID-19. Eur J Epidemiol. 2020; 35 (4): 325–330.

37. Horton R. Offline: COVID-19 is not a pandemic. Lancet 2020; 396: 874.

38. McMahon NE. Understanding COVID-19 through the lens of ‘syndemic vulnerability’: possibilities and challenges. Int J Health Prom and Educ. 2021 (Ahead of print). https://doi.org/10.1080/14635240.2021.1893934

39. Keszei AP, Novak M, Streiner DL. Introduction to health measurement scales. J Psychosom Res. 2010; 68 (4): 319–323.

40. Katz MH. Multivariable analysis. Second edition. Cambridge: Cambridge University Press; 2006. pp. 81–87.

